# The effectiveness of Ear Nose and Throat outreach programs for Aboriginal and Torres Strait Islander Australians: a systematic review

**DOI:** 10.1101/2020.03.04.20031302

**Authors:** Anna Gotis-Graham, Rona Macniven, Kelvin Kong, Kylie Gwynne

**Affiliations:** The University of Sydney; The University of New South Wales; The University of Newcastle

## Abstract

**Background:** Aboriginal and Torres Strait Islander children experience a higher prevalence of ear, nose, and throat (ENT) diseases than non-Indigenous children. Many programs exist that aim to prevent and treat these diseases. Culturally appropriate and timely specialist outreach services may help improve access, service use, and outcomes but there has been a lack of rigorous evaluation of ENT outreach programs to date.

**Objective:** To examine the ability of ENT outreach programs to improve health outcomes among Aboriginal and Torres Strait Islander people

**Methods:** We performed a systematic literature search of nine databases (Medline, CINAHLS, PsychINFO, Embase, Cochrane, Scopus, Global health, Informit Rural health database and Indigenous collection) and grey literature sources for primary studies evaluating ENT outreach services for Aboriginal and Torres Strait Islander Australians. Two authors independently evaluated the eligible articles and extracted relevant information.

**Results:** Of the 506 studies identified, 15 were included in this review. These 15 studies evaluated eight different programs/activities. Studies were heterogeneous in design so a meta-analysis could not be conducted. Seven studies measured health-related outcomes in middle ear or hearing status; six reported overall positive changes one reported no clinically significant improvements. Five programs/activities and their corresponding studies involved Aboriginal and Torres Strait Islander people and organisations in delivery and evaluation, but involvement in program or study design was unclear.

**Conclusion:** While some studies demonstrated improved outcomes, the overall ability of ENT programs/activities to improve health outcomes for Aboriginal and Torres Strait Islander children is unclear. The impact of ENT outreach may be limited by a lack of evidence quality, a lack of coordination of services, and the provision of potentially unsustainable services. Improvements in the quality of evidence, service coordination and sustainability would likely improve health outcomes.

**Strengths and limitations of this study:**

- Studies were identification based on a clearly defined and extensive search strategy based on a priori inclusion and exclusion criteria
- Study appraisal was performed using a relevant tool for mixed methods studies
- The involvement of Aboriginal and Torres Strait Islander people in all aspects of programs and their evaluation was examined

**PROSPERO registration number:** CRD42019134757

## Introduction

Aboriginal and or Torres Strait Islander people represent the oldest continuous living culture in the world, despite ongoing inequities since colonisation [1]. Aboriginal and or Torres Strait Islander children experience a markedly higher prevalence of ear, nose, and throat (ENT) diseases compared to non-Indigenous children [2-4]. Complex historical, cultural and economic factors experienced by Aboriginal and or Torres Strait Islander people since colonisation have interacted with behavioural and biological risk factors to influence this prevalence [2, 4].

The main condition contributing to this prevalence is otitis media, which is the inflammation of the middle ear, usually caused by bacterial and viral pathogens [4]. Compared to non-Indigenous children, Aboriginal and or Torres Strait Islander children tend to experience this preventable and treatable condition at a younger age, more frequently, persistently, and severely, and with more serious complications [3, 5]. Community-based studies have shown the prevalence of otitis media and its complications at up to 73% in those under 12 months of age, and whole communities with otitis media affecting 91% of children [5]. This prevalence is likely perpetuated by socioeconomic factors including poverty, overcrowding, poor nutrition and infrastructure, exposure to cigarette smoke, and limited access to primary health care and treatment [3, 4, 6, 7].

The conductive hearing loss that results from untreated, chronic suppurative otitis media is responsible for the greatest burden of educational, social, and financial sequellae [3]. This is estimated to be 10-30% among Aboriginal and Torres Strait Islander children, well above the World Health Organisations cut off for ‘a massive public health problem requiring urgent attention’, which they quote as above 4% [3]. Hearing loss contributes to early learning difficulties including speech delays with resulting low self-esteem, poorer education outcomes [8, 9] and a significant economic burden [4]. This significant impact affects their long-term quality of life and life opportunities.

The majority of otitis media is managed in primary healthcare with referral to ENT specialists for assessment and surgical interventions where appropriate [10]. In Australia, referral to ENT specialists care is complex and varies across jurisdictions, with limited access to public ENT clinics [11]. However, in rural and remote settings, Aboriginal and or Torres Strait Islander children face wait times that are longer than recommended for audiology testing and ENT services, with a higher likelihood that these services are unavailable. While the practice of most ENT surgeons in Australia is largely confined to metropolitan areas, fewer participate in outreach clinics to rural and remote areas [11]. To access ENT specialist services, patients are generally required to overcome barriers including travel, culturally inappropriate services, and unfamiliar health-system processes [12, 13]. Furthermore, the current system fails to routinely deliver care that aligns with government guidelines [14], nor provide culturally safe and accessible clinical pathways. Outreach services mobilise the expertise of health care teams and individual practitioners away from their usual place of work, generally to an underserviced area. This may take the form of traditional fly-in-fly-out services, or newer remote telemedicine enabled services. These services may be in a unique position to combat the challenges faced by the current system, with evidence for improved access, outcomes, service use, and less disruption to patient and family life when employed with well-functioning primary care services [15-18]. The aim is to provide a service that is truly accessible by the Aboriginal and or Torres Strait Islander community. While the role and benefits of outreach services are generally well recognised, rigorous evaluation of existing outreach programs is lacking, including those pertaining to ENT specialties, and as such, little is known about the impact and outcomes of such programs [19, 20]. The result is the implementation of programs without sufficient planning or evidence base [2]. This review aims primarily to examine the ability of ENT outreach programs to improving Aboriginal and Torres Strait Islander health outcomes, and secondarily to elucidate factors predicting success, and barriers to success of such programs.

## Methods

### Study design

This study is a systematic review of peer-reviewed and grey literature and is reported using the Preferred Reporting Items for Systematic Reviews and Meta-Analysis (PRISMA) statement guidelines [21]. The study was registered with PROSPERO International prospective register of systematic reviews (CRD42019134757) [22].

### Eligibility criteria

This review sought to identify studies that supplied ENT outreach services to Aboriginal and or Torres Strait Islander Australians and provided data to evaluate their aims. Studies were included according to the following criteria

- Population: all or predominately Aboriginal or Torres Strait Islander Australian participants
- Intervention: ENT outreach services including but not limited to screening, management, or rehabilitation of ENT disease
- Comparator: as determined by the nominated study
- Outcome: as determined by the nominated study
- Study design: all study types

The search was limited to English language studies published between 2000-2018 inclusive. Studies were excluded if they did not provide primary data; or if they were descriptive only or aimed to identify the incidence or prevalence of disease without intervention or referral for subsequent treatment. The former criteria are applied as it necessarily precludes an objective evaluation, while the latter is applied as programs without intervention or referral pathways do not improve the health of Aboriginal and or Torres Strait Islander Australians [23].

### Search strategy

Nine online databases (Medline, CINAHLS, PsychINFO, Embase, Cochrane, Scopus, Global health, Informit Rural health database and Indigenous collection) were searched for published articles between December 2018 and January 2019. An example search is provided in Appendix 1. A grey literature search of relevant government and non-government websites was conducted including the Rural Doctors Network, Australian Indigenous HealthInfoNet, Australian Institute of Health and Welfare, Rural Health, The Lowitja Institute, Australian state and national government health departments. Where conference papers were identified or a study’s full text unavailable, contact was made with authors to source original data. The reference lists of included studies, and other systematic reviews identified in the literature search, were screened for additional eligible studies.

### Study selection process

Following duplicate removal, the first and second author screened a random sample of 25% to identify studies congruent with the inclusion criteria. Discrepancies were resolved by consensus between the two authors and the first author screened the remaining studies. This process was repeated for articles identified in the grey literature. The first and second authors independently assessed full texts for eligibility. Discrepancies were resolved by consensus, generating a final list of studies for inclusion.

### Data extraction and synthesis

Data from included studies were extracted according to program characteristics (Program name and aims, operating years, State/Territory, Area, Setting, Disease focus, Indigenous capacity building) and evaluation characteristics (study aim, study type, outcome measures, participant number and age, main findings). The first and second authors independently extracted a sample of texts and reviewed results, with discrepancies identified and resolved by consensus. The first author extracted the remaining texts according to consensus. Where studies also reported on outreach services of other specialties, only ENT-specific outcomes were included in this report. Studies were analysed in a qualitative synthesis. Meta-analysis was deemed inappropriate due to the small study sample sizes, mixed methods study inclusion, and the heterogeneity of the study designs and outcome.

### Risk of bias

Risk of bias was assessed using the Mixed Methods Assessment Tool (MMAT) [24]. The first and second authors independently assessed a sample of studies and reviewed results with discrepancies identified and resolved by consensus. The first author assessed the remaining texts according to consensus.

## Results

### Study selection

The database search, grey literature, and hand search identified 930, 34 articles, and 5 studies respectively, with 506 remaining following duplicate removal. Of the 506 studies that were screened, 434 were excluded and 72 underwent full-text review. A further 54 were excluded ultimately leaving 15 articles included in the review (Figure 1).

**Figure 1.**
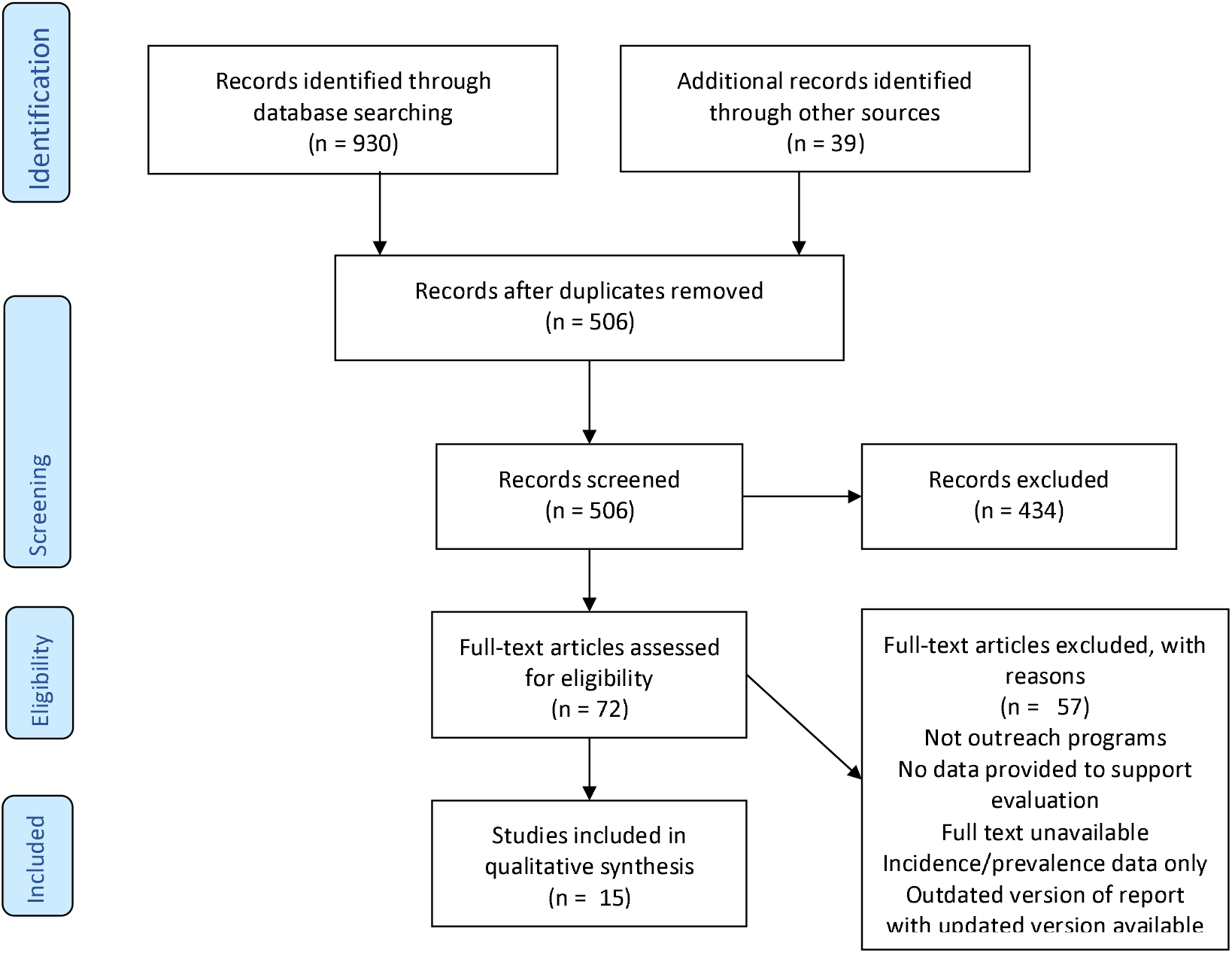
PRISMA flow chart.

### Program characteristics

Table 1 provides the program/activity characteristics of included studies

**Table 1:**
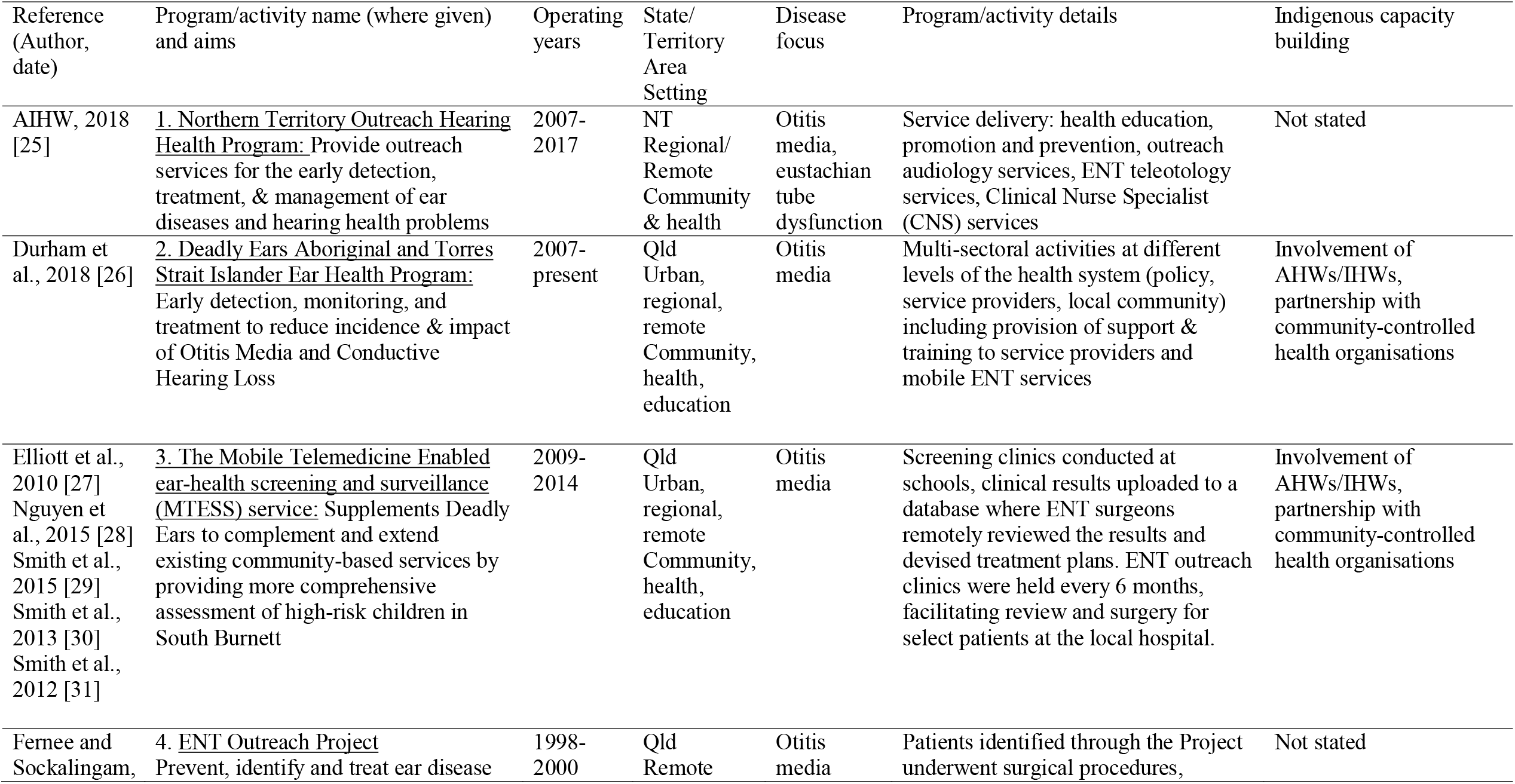
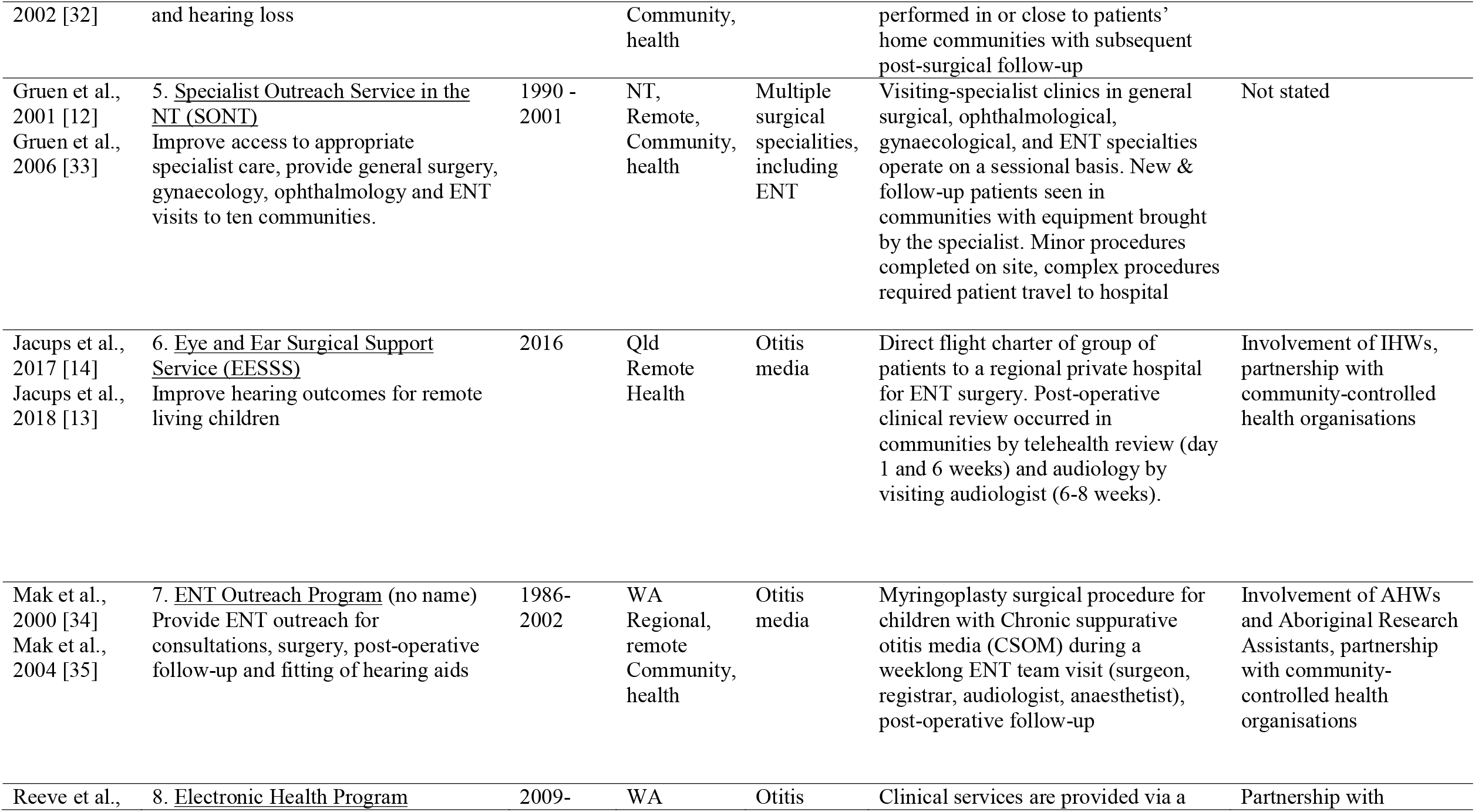
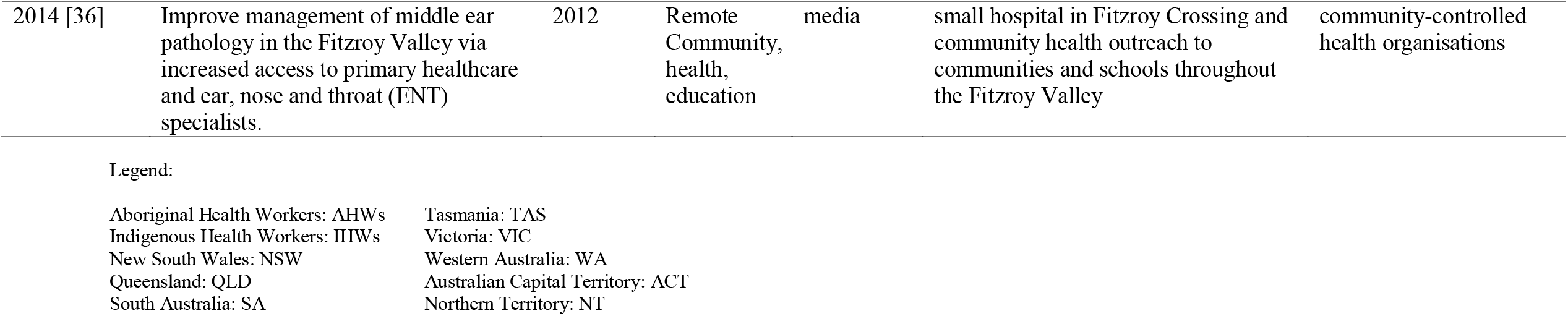
Program characteristics of included studies

| Reference<br>(Author,<br>date) | Program/activity name (where given)<br>and aims | Operating<br>years | State/<br>Territory<br>Area<br>Setting | Disease<br>focus | Program/activity details | Indigenous capacity<br>building |
| --- | --- | --- | --- | --- | --- | --- |
| AIHW, 2018<br>[25] | <u>1. Northern Territory Outreach Hearing Health Program:</u> Provide outreach services for the early detection, treatment, & management of ear diseases and hearing health problems | 2007-2017 | NT<br>Regional/<br>Remote<br>Community<br>& health | Otitis<br>media,<br>eustachian<br>tube<br>dysfunction | Service delivery: health education, promotion and prevention, outreach audiology services, ENT teleotology services, Clinical Nurse Specialist (CNS) services | Not stated |
| Durham et<br>al., 2018 [26] | <u>2. Deadly Ears Aboriginal and Torres Strait Islander Ear Health Program:</u> Early detection, monitoring, and treatment to reduce incidence & impact of Otitis Media and Conductive Hearing Loss | 2007-present | Qld<br>Urban,<br>regional,<br>remote<br>Community,<br>health,<br>education | Otitis<br>media | Multi-sectoral activities at different levels of the health system (policy, service providers, local community) including provision of support & training to service providers and mobile ENT services | Involvement of<br>AHWs/IHWs,<br>partnership with<br>community-controlled<br>health organisations |
| Elliott et al.,<br>2010 [27]<br>Nguyen et<br>al., 2015 [28]<br>Smith et al.,<br>2015 [29]<br>Smith et al.,<br>2013 [30]<br>Smith et al.,<br>2012 [31] | <u>3. The Mobile Telemedicine Enabled ear-health screening and surveillance (MTESS) service:</u> Supplements Deadly Ears to complement and extend existing community-based services by providing more comprehensive assessment of high-risk children in South Burnett | 2009-2014 | Qld<br>Urban,<br>regional,<br>remote<br>Community,<br>health,<br>education | Otitis<br>media | Screening clinics conducted at schools, clinical results uploaded to a database where ENT surgeons remotely reviewed the results and devised treatment plans. ENT outreach clinics were held every 6 months, facilitating review and surgery for select patients at the local hospital. | Involvement of<br>AHWs/IHWs,<br>partnership with<br>community-controlled<br>health organisations |
| Ferneer and<br>Sockalingam, | <u>4. ENT Outreach Project</u><br>Prevent, identify and treat ear disease | 1998-2000 | Qld<br>Remote | Otitis<br>media | Patients identified through the Project underwent surgical procedures, | Not stated |
| 2002 [32] | and hearing loss |  | Community, health |  | performed in or close to patients' home communities with subsequent post-surgical follow-up |  |
| Gruen et al., 2001 [12]<br>Gruen et al., 2006 [33] | 5. <u>Specialist Outreach Service in the NT (SONT)</u><br>Improve access to appropriate specialist care, provide general surgery, gynaecology, ophthalmology and ENT visits to ten communities. | 1990 - 2001 | NT, Remote, Community, health | Multiple surgical specialities, including ENT | Visiting-specialist clinics in general surgical, ophthalmological, gynaecological, and ENT specialties operate on a sessional basis. New & follow-up patients seen in communities with equipment brought by the specialist. Minor procedures completed on site, complex procedures required patient travel to hospital | Not stated |
| Jacups et al., 2017 [14]<br>Jacups et al., 2018 [13] | 6. <u>Eye and Ear Surgical Support Service (EESSS)</u><br>Improve hearing outcomes for remote living children | 2016 | Qld Remote Health | Otitis media | Direct flight charter of group of patients to a regional private hospital for ENT surgery. Post-operative clinical review occurred in communities by telehealth review (day 1 and 6 weeks) and audiology by visiting audiologist (6-8 weeks). | Involvement of IHWs, partnership with community-controlled health organisations |
| Mak et al., 2000 [34]<br>Mak et al., 2004 [35] | 7. <u>ENT Outreach Program</u> (no name)<br>Provide ENT outreach for consultations, surgery, post-operative follow-up and fitting of hearing aids | 1986-2002 | WA Regional, remote Community, health | Otitis media | Myringoplasty surgical procedure for children with Chronic suppurative otitis media (CSOM) during a weeklong ENT team visit (surgeon, registrar, audiologist, anaesthetist), post-operative follow-up | Involvement of AHWs and Aboriginal Research Assistants, partnership with community-controlled health organisations |
| Reeve et al., | 8. <u>Electronic Health Program</u> | 2009- | WA | Otitis | Clinical services are provided via a | Partnership with |
| 2014 [36] | Improve management of middle ear pathology in the Fitzroy Valley via increased access to primary healthcare and ear, nose and throat (ENT) specialists. | 2012 | Remote Community, health, education | media | small hospital in Fitzroy Crossing and community health outreach to communities and schools throughout the Fitzroy Valley | community-controlled health organisations |

Legend:
|  |  |
| --- | --- |
| Aboriginal Health Workers: AHWs | Tasmania: TAS |
| Indigenous Health Workers: IHWs | Victoria: VIC |
| New South Wales: NSW | Western Australia: WA |
| Queensland: QLD | Australian Capital Territory: ACT |
| South Australia: SA | Northern Territory: NT |

Publication dates of included studies span 2000 – 2018, with most published since 2010. Most studies evaluated programs for greater than twelve months. Those evaluated for less than twelve months represented feasibility studies [27], or a program of two months’ duration [13, 14]. All programs focused on the management of ear disease or sequellae, predominately otitis media.

Overall, eight programs were represented by 15 studies. The National Partnership on Northern Territory (NT) Remote Aboriginal Investment activities is represented by the NT Outreach Hearing Health Program (HHP) and evaluated by the Australian Government [25]. Queensland (Qld) Deadly Ears Aboriginal and Torres Strait Islander Ear Health Program was evaluated in one study that examined the alignment between the program framework and systems thinking concepts [26]. A supplement to Deadly Ears, The Mobile Telemedicine Enabled ear-health screening and surveillance (MTESS) service was evaluated by five studies [27-31]. These studies evaluated service feasibility in terms of community acceptance, integration, and practical feasibility [27]; program outcomes after three years [31]; changes in hospital referral patterns [30]; changes in screening patterns [29]; and the cost-effectiveness [28]. A new service model within the Eye and Ear Surgical Support Service (EESSS) via CheckUp Australia, where patients were chartered to a regional basin for ENT services, was evaluated by two studies [13, 14]. The first evaluated the clinical and hearing outcomes of participants following the intervention [14] while the second compared model cost with existing services and alternative approaches [13]. The Specialist Outreach Service in the NT (SONT) was evaluated by two studies across different time points [12, 33]. The first examined program effectiveness and identify barriers to specialist access [12], while the second assessed program effects on access, referral patterns, and care outcomes [33]. The outcomes of ENT surgical interventions as part of an unspecified outreach program were evaluated by two studies at two different time points [34, 35]. The remaining two programs were an ENT Outreach Project [32] and an Electronic Health Program, [36] which were evaluated by single studies.

Two programs took place in the NT [12, 25, 33], four in Qld [13, 14, 26-32] and two in Western Australia (WA) [34-36]. Settings varied with two programs providing school or community based screening with outreach follow up [27-31, 36], four programs facilitating visiting specialists to local communities providing clinics or surgical intervention [12-14, 32-35], while two were state-based programs and frameworks delivered services across multiple settings [25, 26].

Interventions varied in nature with two programs providing ear screening services with telemedicine enabled ENT follow up [27-31, 36], three programs providing surgical interventions for the management of ear disease [13, 14, 32, 34, 35], two providing multiple services as part of state-wide programs [25, 26] and one program providing fly-in-fly-out ENT specialist clinics [12, 33].

There was varying involvement of Aboriginal and Torres Strait Islander people in program design, service delivery or evaluation with only studies associated with the MTESS reporting ongoing involvement in all three stages [27-31]. Five programs had stated involvement or partnerships with Aboriginal Community Controlled Health Services (ACCHSs) engaged Aboriginal Health Workers (AHWs) or Indigenous Health Workers (IHW) in program/activity or evaluation [13, 14, 26, 29-31, 34-36].

Table 2 provides details of study evaluation characteristics.

**Table 2:**
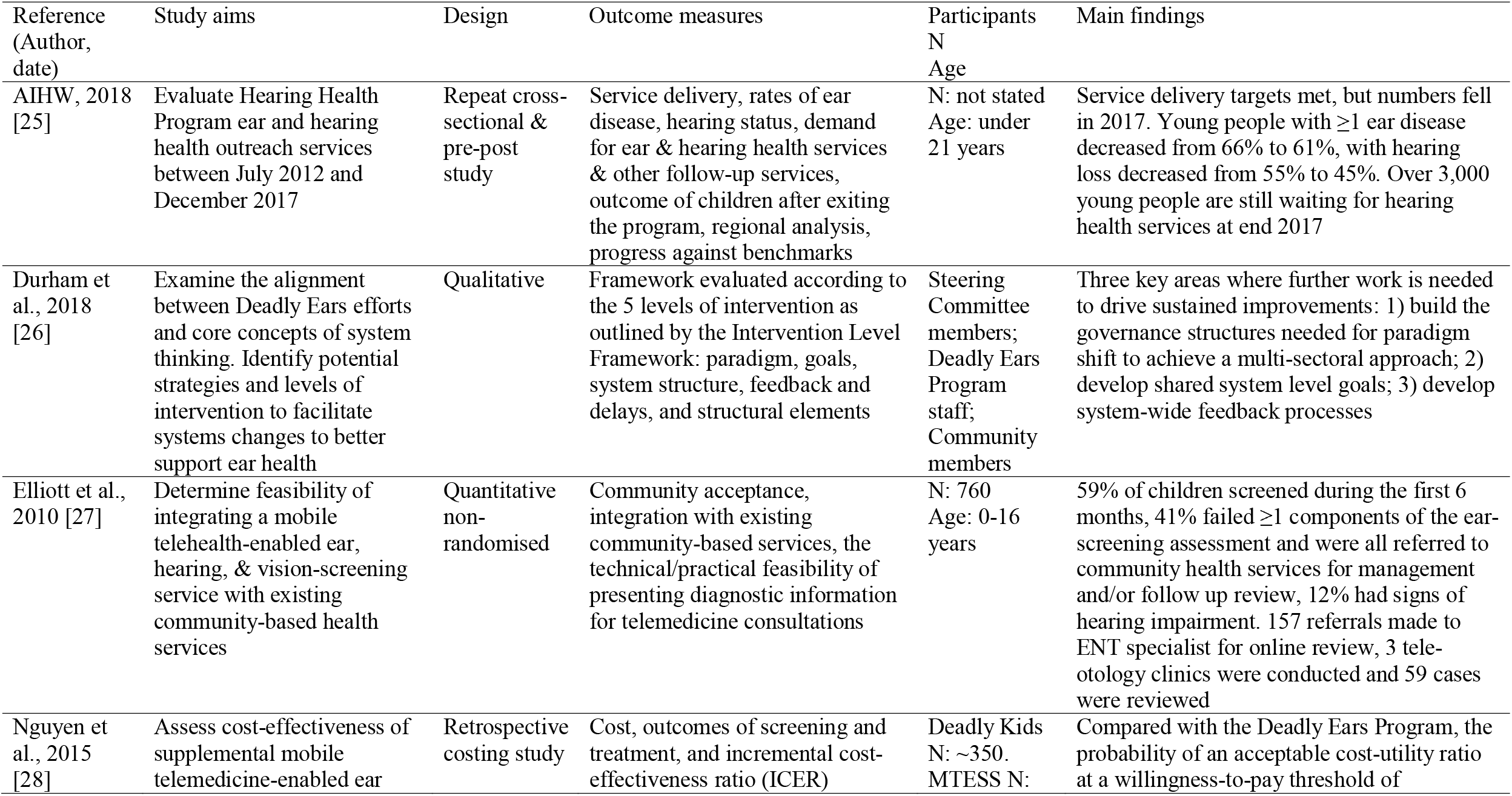
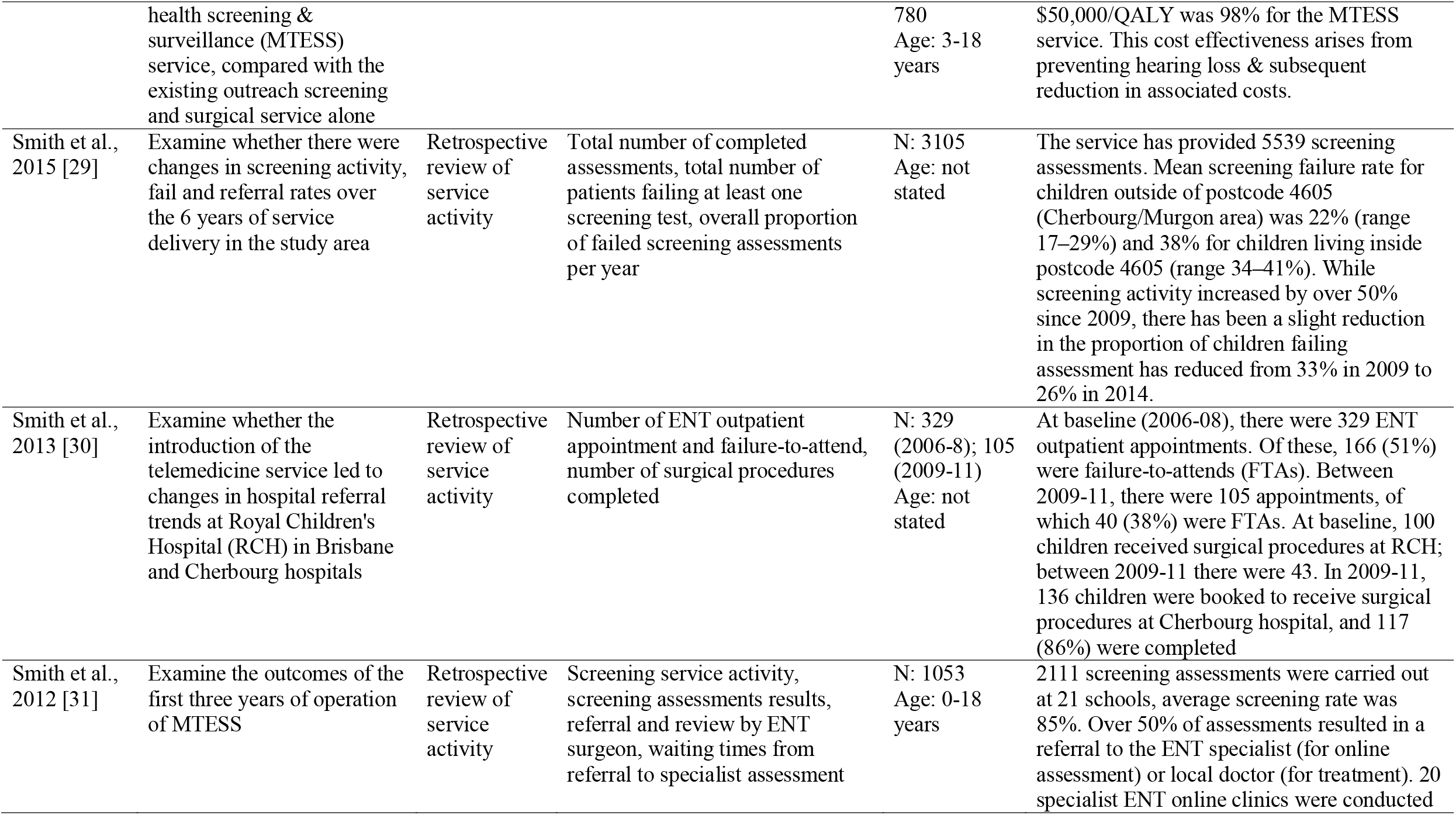
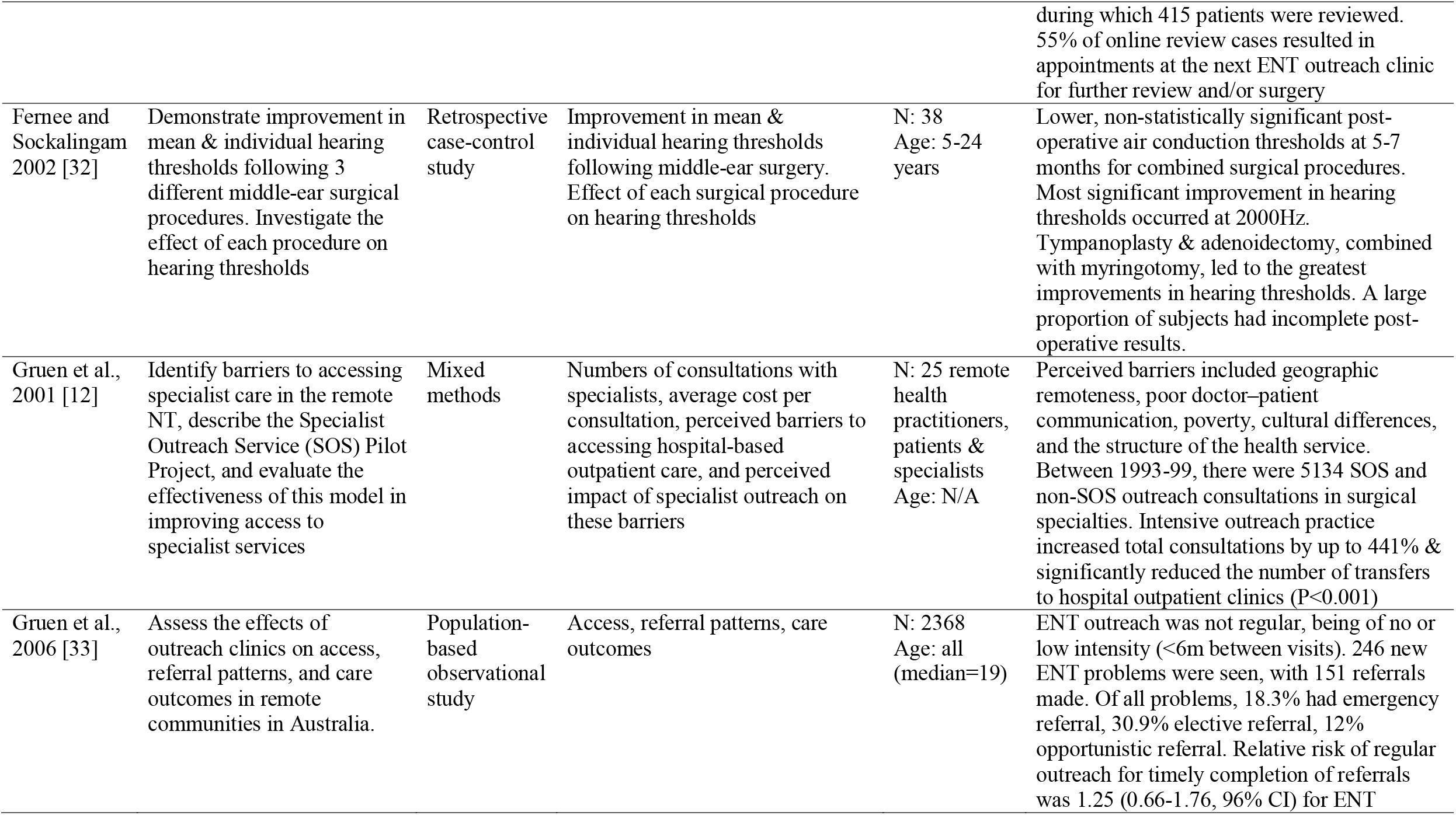
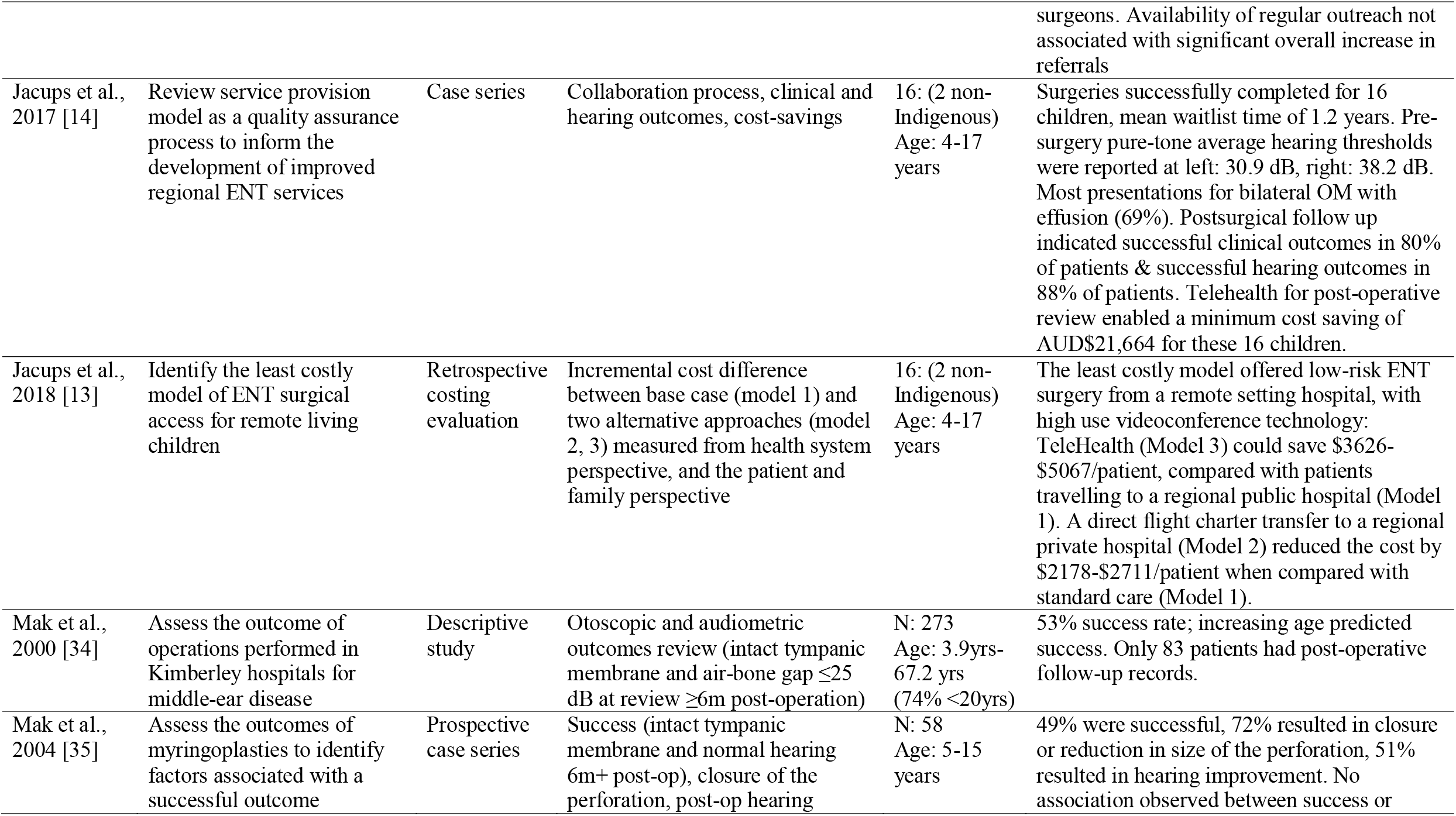
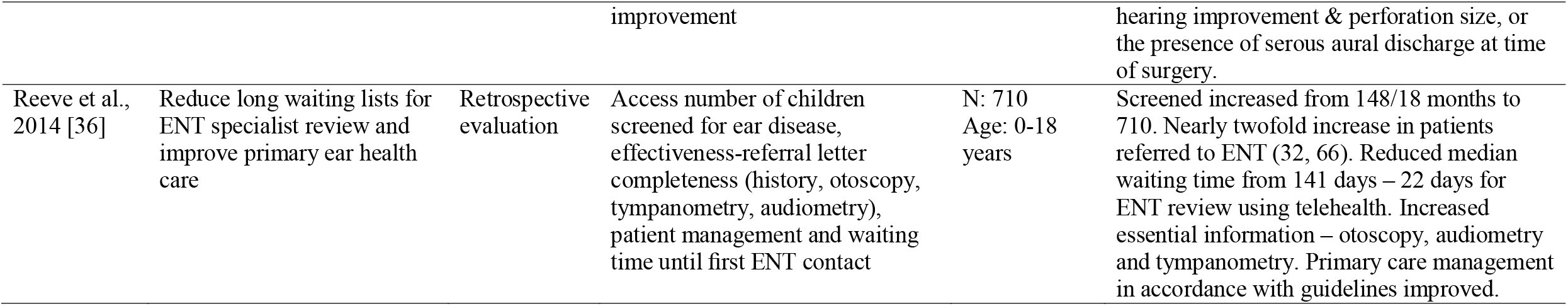
Evaluation characteristics of included studies

There was heterogeneity in study aims where six studies aimed to evaluate intervention effects on health-related outcomes [14, 25, 29, 32, 34, 35], four studies aimed to evaluate program effectiveness on accessibility and referral pathways [30, 31, 33, 36], two aimed to conduct cost analyses of delivered and proposed models of care [13, 28], two aimed to assess program feasibility or identify barriers to care [12, 27], and one aimed to examine the alignment between a program framework and the concepts of systems thinking [26]. This heterogeneity is also reflected in study types with five retrospective reviews of service activity [29-31, 34, 36], two retrospective costing studies [14, 28], two case series [14, 35], three mixed method analyses [12, 26], one repeat cross-sectional study [25], one quantitative non-randomised study [27], one retrospective case-control study [32] and one population based observational study [33]. Outcome measures were equally heterogeneous with data based on qualitative and quantitative methods including, but not limited to screening rates, hearing thresholds, middle ear and tympanic membrane status, referral rates, waiting time for services, appropriateness of primary care management, and cost difference.

Most outcome measures provided data that supported, but did not directly allow for, the rigorous evaluation of broader program aims. While studies such as cost analyses [13, 28] did not directly link to program aims, they provide important information regarding feasibility and sustainability of program implementation. Outreach programs appeared to have increased service delivery levels, but this was frequently reported without reference to baseline rates [26, 33] making the change difficult to quantify. Three programs reported decreased waiting times for ENT review or surgery [14, 31, 36], but these remained lengthy for a number of children in the NT HHP [25].

Of the eight programs, six were evaluated by studies that measured health-related outcomes in either middle ear or hearing status. Five studies reported overall positive changes in middle ear and/or hearing health [14, 25, 29, 34, 35] and one reported no clinically significant improvements [32]. The state-wide NT HHP program reported decreasing the need for follow up, medical or surgical treatment [25] while local services increased the number of follow up referrals within three programs [30, 31, 36]. Programs did not increase the demand on outpatient services [30, 33].

### Risk of bias

All papers were empirical studies and were suitable for appraisal using the MMAT (Appendix 2). Overall, the quality of studies was generally poor with only two of the 15 studies scoring ‘yes’ on all five MMAT measures.

## Discussion

This review examined the ability of ENT outreach programs to improve health outcomes among Aboriginal and Torres Strait Islander children and included 15 studies of eight programs. We found that while study outcome measures were linked to program aims, these links could be peripheral or did not provide sufficiently rigorous data to evaluate programs. Study characteristics varied widely but overall, positive changes were seen in middle ear and/or hearing health in five of the eight programs [14, 25, 29, 34, 35]. These results should be interpreted carefully as all measures of service delivery, referral rates, attendance rates, wait times, were difficult to contextualise given a lack of baseline data and inter-study variation in the methods and clinical thresholds used to monitor changes.

Despite these limitations, these six programs appear to produce positive results in the communities which they are delivered. However, the limited quantity and quality of evidence, a lack of coordination of programs, and the appropriateness and acceptability of services is likely contributing to the ongoing burden of ear disease in Aboriginal and or Torres Strait Islander children [2]. Although over 50 current or recent ear health programs exist across Australia [37], we found a paucity of literature evaluating their programs. This is consistent with a recent review of physical activity programs for Aboriginal and Torres Strait Islander people that found that while many programs existed, few were comprehensively evaluated [38]. Among the included studies, there was marked heterogeneity in the setting and nature of interventions and evaluation, including their outcome measures. A lack of standardised systems for monitoring changes in incidence and prevalence of ear disease limits the ability to measure and attribute changes in disease states to the actions of a program [26, 39]. However, regular evaluation in the form of continuous quality improvement frameworks have been shown to improve the quality of healthcare for Aboriginal and or Torres Strait Islander children as well as health outcomes in other areas including antenatal care, immunisations, smoking, alcohol consumption, diabetes, cardiovascular disease, and cervical screening [40, 41]. Several ear health indicators that are potentially extractable from electronic health records have been recommended [41]. Though these indicators have only been validated in the primary health setting so far, there is potential for their use in the ongoing evaluation of ENT outreach programs.

Sustainable outreach benefits in disease prevention, treatment, and management may occur with coordinated service delivery [2, 42]. We are limited in our ability to draw conclusions regarding the coordination of all Australian ENT services as many programs do not provide evaluation data that could be included in this systematic review. The included programs took place across three states, Qld, the NT and WA, in multiple settings. Services were delivered as part of, or in association with numerous programs with little to no evidence of interaction or coordination between these programs in terms of aims, service delivery, coverage, or funding bodies. Effective outreach programs require efficient integration of incoming ENT services with existing primary health care services and the broader community [17]. One program, the MTESS, reported integration with the community through the local AHW and close alignment with primary care services to be important factors in success [29, 30] and this is recommended to strengthen future program delivery.

There is currently a discordance between service delivery and burden of disease [15]. A significant barrier to coordination is the lack of population level data detailing the epidemiology of ear disease in Aboriginal and or Torres Strait Islander children [26, 39] as strategic delivery of services is limited when need cannot be directly pinpointed. A national outreach service register has been suggested as a way of identifying areas of over or under supply [17]. The MTESS reports the probability of service uptake in areas was directly linked to the provision of services [28], reiterated by the HHP where audiology, ENT, and clinical nurse specialist service numbers dropped following a shortage of available specialists [25]. The result is ad-hoc service delivery contributing to a lack of coordination, inequity and unsustainable service delivery [16-18, 42]. Finally, while outreach programs play a role in improving the health of Aboriginal and or Torres Strait Islander children, they form only one piece of the puzzle. Coordination is required between multiple sectors to effectively address the socioeconomic and historical aetiology of ear disease. While the DEDKDC Framework prioritised multi-sector collaboration and coordination, there was little evidence of these activities [26].

Outreach programs are often large and multi-faceted, leading to complexities in evaluation. Studies that are fragmented from program aims impede the development of program learnings and limits the ability of past programs to critically inform the development of future programs. The scarcity of program evaluation hinders a global assessment of factors predicting success, or barriers to success of ENT outreach programs, as originally planned in this review. However, factors known to impact on the success of outreach programs are regular and predictable service, and communication with and accountability to the community [18]. The frequency and regularity of outreach events in programs included in this review were largely unclear and a lack of accountability leads to irregularity and unpredictability, creating issues with delayed follow up and inadequate support [18].

Furthermore, this review revealed a deficiency in collaboration with communities in planning, service delivery, and evaluation of included programs, indicating a lack of communication with and accountability to the community. This finding is consistent with the literature where the status quo sees services for Aboriginal and or Torres Strait Islander peoples developed without their input [43]. Acceptable healthcare delivery relies on effective collaboration, which necessitates the genuine involvement of Aboriginal and Torres Strait Islander people and valuing of traditional practices [44, 45]. MTESS service highlighted the importance of this process in their ability to sustain integration of the service with primary healthcare and the deliver ongoing convenient and timely services [29]. Furthermore, activities run under the DEDKDC Framework were reported to have greater attendance where AHWs were present [46]. Other recommendations for sustainability include an adequate primary care and specialist base, a multidisciplinary framework centred in primary care, funding and coordination that recognises responsibilities primary, secondary and tertiary care and regular evaluation. [18].

There is a strengthened resolve when Aboriginal and Torres Strait Islander people are involved in the planning, running and evaluation of ENT outreach clinics. Codesign and capacity building must be part of any outreach program and continual re-evaluation is strongly recommended. There is also a need to ensure services are complemented by programs that add value and with translational research outcomes that support communities. Working solutions backed by evidence and community benefits need to be published and supported broadly to apply to local conditions.

## Conclusion

This review discovered a paucity of evaluation of ENT outreach programs for Aboriginal and Torres Strait Islander children. Fifteen evaluations of eight programs were identified that were heterogeneous in study design and of variable methodological quality. While individual studies reflected positive outcomes of programs, including positive changes in middle ear and/or hearing health from six programs, the ability of these programs to improve the overall ear-health status of Aboriginal children remains unclear. These findings suggest that the effectiveness of ENT outreach programs may be limited by a lack of coordination of services and the provision of potentially unsustainable services. There were also low levels of involvement of Aboriginal and Torres Strait Islander people in program and evaluation design and delivery and we recommend greater involvement in all future program and evaluation aspects to strengthen their impact and outcomes.

## Data Availability

All data relevant to the study are included in the article or uploaded as supplementary information.

## References

1. Paradies, Y., Colonisation, racism and indigenous health. Journal of Population Research, 2016. 33(1): p. 83–96.

2. Closing the Gap Clearinghouse (AIHW & AIFS), Ear disease in Aboriginal and Torres Strait Islander children, C.t.G. Clearinghouse, Editor. 2014, AIHW & AIFS: Canberra & Melbourne.

3. Jervis-Bardy, J., L. Sanchez, and A.S. Carney, Otitis media in indigenous australian children: Review of epidemiology and risk factors. Journal of Laryngology and Otology, 2014. 128(S1): p. S16–S27.

4. Burns, J. and N. Thomson, Review of ear health and hearing among Indigenous Australians. Australian Indigenous HealthBulletin, 2013. 13(4).

5. Morris, P.S., et al., Otitis media in young Aboriginal children from remote communities in Northern and Central Australia: a cross-sectional survey. BMC pediatrics, 2005. 5 (27).

6. Jacoby, P.A., et al., The effect of passive smoking on the risk of otitis media in Aboriginal and non-Aboriginal children in the Kalgoorlie-Boulder region of Western Australia. Medical Journal of Australia, 2008. 188(10): p. 599–603.

7. Leach, A.J., et al., Bacterial colonization of the nasopharynx predicts very early onset and persistence of otitis media in Australian Aboriginal infants. Pediatric Infectious Disease Journal, 1994. 13(11): p. 983–989.

8. Kong, K. and H.L. Coates, Natural history, definitions, risk factors and burden of otitis media. Medical Journal of Australia, 2009. 191(S9): p. S39–S43.

9. Brown, M., N. Wigg, and K. Turner, Ear disease and indigenous kids: Tackling the silent epidemic. Journal of Paediatrics and Child Health, 2011. 47(S2): p. S18–S19.

10. Couzos, S., S. Metcalf, and R.B. Murray, Systematic Review of Existing Evidence and Primary Care Guidelines on the Management of Otitis Media in Aboriginal and Torres Strait Islander Populations. 2001, Commonwealth Department of Health and Aged Care: Canberra, ACT, Australia.

11. Shein, G., et al., The O.P.E.N. Survey: outreach projects in Ear, Nose and Throat (ENT) in New South Wales. Australian Journal of Otolaryngology, 2019. 2.

12. Gruen, R.L., et al., Improving access to specialist care for remote Aboriginal communities: evaluation of a specialist outreach service. Medical Journal of Australia, 2001. 174(10): p. 507–511.

13. Jacups, S.P., I. Kinchin, and K.M. McConnon, Ear, nose, and throat surgical access for remote living Indigenous children: What is the least costly model? Journal of Evaluation in Clinical Practice, 2018. 24(6): p. 1330–1338.

14. Jacups, S.P., et al., An innovative approach to improve ear, nose and throat surgical access for remote living Cape York Indigenous children. International Journal of Pediatric Otorhinolaryngology, 2017. 100: p. 225–231.

15. Gunasekera, H., et al., Otitis media in Aboriginal children: The discordance between burden of illness and access to services in rural/remote and urban Australia. Journal of Paediatrics and Child Health, 2009. 45(7-8): p. 425–430.

16. Gruen, R. and R. Bailie, Specialist clinics in remote Australian Aboriginal communities: where rock art meets rocket science. Journal of Health Services Research and Policy, 2004. 9(S2): p. S56–S62.

17. O’Sullivan, B.G., C.M. Joyce, and M.R. McGrail, Adoption, implementation and prioritization of specialist outreach policy in Australia: A national perspective. Bulletin of the World Health Organization, 2014. 92(7): p. 512–519.

18. Gruen, R.L., T.S. Weeramanthri, and R.S. Bailie, Outreach and improved access to specialist services for Indigenous people in remote Australia: the requirements for sustainability. Journal of Epidemiology and Community Health, 2002. 56(7): p. 517–521.

19. Coates, H.L., et al., Otitis media in Aboriginal children: tackling a major health problem. Medical Journal of Australia, 2002. 177(4): p. 177–178.

20. Mbuzi, V., P. Fulbrook, and M. Jessup, Effectiveness of programs to promote cardiovascular health of Indigenous Australians: A systematic review. International Journal for Equity in Health, 2018. 17(153).

21. Moher, D., et al., Preferred reporting items for systematic reviews and meta-analyses: the PRISMA statement. Ann Intern Med, 2009. 151.

22. Gotis-Graham, A., R. MacNiven, and K. Gwynne. PROSPERO: The effectiveness of ENT outreach interventions on the health of Indigenous Australians: a systematic review. 2019 25.11.2019]; Available from: https://www.crd.york.ac.uk/PROSPERO/display_record.php?RecordID=134757.

23. Huntley, P., B. Woods, and S. Rudge, Healthy Ears, Happy Kids: a new approach to Aboriginal child ear health in NSW, in New South Wales Public Health Bulletin. 2012: Victoria. p. 60–61.

24. Pace, R., et al., Testing the reliability and efficiency of the pilot Mixed Methods Appraisal Tool (MMAT) for systematic mixed studies review. International Journal of Nursing Studies, 2012. 49(1): p. 47–53.

25. AIHW, Northern Territory Outreach Hearing Health Program: July 2012 to December 2017. 2018, AIHW: Canberra.

26. Durham, J., et al., Using systems thinking and the Intervention Level Framework to analyse public health planning for complex problems: Otitis media in Aboriginal and Torres Strait Islander children. PLoS ONE, 2018. 13(3): p. e0194275.

27. Elliott, G., et al., The Feasibility of a Community-Based Mobile Telehealth Screening Service for Aboriginal and Torres Strait Islander Children in Australia. Telemedicine and e-Health, 2010. 16(9): p. 950–956.

28. Nguyen, K.H., et al., Cost-Effectiveness Analysis of a Mobile Ear Screening and Surveillance Service versus an Outreach Screening, Surveillance and Surgical Service for Indigenous Children in Australia. PLoS ONE, 2015. 10 (9): p. e0138369.

29. Smith, A.C., et al., Monitoring ear health through a telemedicine-supported health screening service in Queensland. Journal of Telemedicine & Telecare, 2015. 21(8): p. 427–430.

30. Smith, A.C., et al., Changes in paediatric hospital ENT service utilisation following the implementation of a mobile, indigenous health screening service. Journal of Telemedicine & Telecare, 2013. 19(7): p. 397–400.

31. Smith, A.C., et al., A mobile telemedicine-enabled ear screening service for Indigenous children in Queensland: activity and outcomes in the first three years. Journal of Telemedicine & Telecare, 2012. 18(8): p. 485–489.

32. Fernee, B. and R. Sockalingam, Outcomes of ENT surgery for middle-ear disease in aboriginal populations living in remote communities: A comparison between pre and post operative audiometric results. Australian Journal of Otolaryngology, 2002. 5(1): p. 6–13.

33. Gruen, R.L., et al., Specialist outreach to isolated and disadvantaged communities: a population-based study. Lancet, 2006. 368: p. 130–138.

34. Mak, D., et al., Middle-ear disease in remote Aboriginal Australia: A field assessment of surgical outcomes. Journal of Laryngology and Otology, 2000. 114(1): p. 26–32.

35. Mak, D., et al., Outcomes of myringoplasty in Australian aboriginal children and factors associated with success: A prospective case series. Clinical Otolaryngology and Allied Sciences, 2004. 29(6): p. 606–611.

36. Reeve, C., et al., Evaluation of an ear health pathway in remote communities: Improvements in ear health access. Australian Journal of Rural Health, 2014. 22(3): p. 127–132.

37. HealthInfoNet. Programs - Ear Health - Australian Indigenous HealthInfoNet. 2018 01.12.18]; Available from: https://healthinfonet.ecu.edu.au/learn/health-topics/ear-health/programs-and-projects/?&topicid=0&topic=all&pagenum=2&sorter=1.

38. Macniven, R., et al., A snapshot of physical activity programs targeting Aboriginal and Torres Strait Islander people in Australia. Health Promotion Journal of Australia, 2017. 28(3): p. 185–206.

39. Durham, J., L. Schubert, and L. Vaughan, Deadly Ears Deadly Kids Deadly Communities framework evaluation report. 2015, Queensland Health: Brisbane.

40. McAullay, D., et al., Sustained participation in annual continuous quality improvement activities improves quality of care for Aboriginal and Torres Strait Islander children. Journal of Paediatrics and Child Health, 2018. 54(2): p. 132–140.

41. Sibthorpe, B., et al., Indicators for continuous quality improvement for otitis media in primary health care for Aboriginal and Torres Strait Islander children. Australian Journal of Primary Health, 2017. 23(1): p. 1–9.

42. O’Sullivan, B.G., C.M. Joyce, and M.R. McGrail, Rural outreach by specialist doctors in Australia: a national cross-sectional study of supply and distribution. Human Resources for Health, 2014. 12(50).

43. McIntosh, K. and L. Oke, Weenthunga meaning hearing and listening. International Journal of Stroke, 2010. 1): p. 45.

44. Ware, V.A., Improving the accessibility of health services in urban and regional settings for Indigenous people, C.t.G. Clearinghouse, Editor. 2013, AIHW & AIFS: Canberra & Melbourne.

45. Lucero, E., From tradition to evidence: Decolonization of the evidence-based practice system. Journal of Psychoactive Drugs, 2011. 43(4): p. 319–324.

46. Durham, J., L. Vaughan, and L. Schubert, Towards a programme theory in the application of systems thinking to complex public health issues. Tropical Medicine and International Health, 2015. 1): p. 99–100.

